# Refining LLMs Outputs with Iterative Consensus Ensemble (ICE)

**DOI:** 10.1101/2024.12.25.24319629

**Authors:** Mahmud Omar, Benjamin S Glicksberg, Girish N Nadkarni, Eyal Klang

**Author notes:** **Corresponding author:** Mahmud Omar M.D., Institutional.

## Abstract

Large language models (LLMs) show promising accuracy on challenging tasks, including medical question answering. Yet, direct gains from model upgrades can plateau, and reliability issues persist. We introduce Iterative Consensus Ensemble (ICE), a proof-of-concept framework that refines answers through iterative reasoning and feedback among multiple LLMs. This ensemble method encourages diverse models to scrutinize each other’s outputs, converging on a consensus solution. We tested ICE on four different datasets. These included over 4,000 multiple-choice questions drawn from a newly curated primary care exam set, established medical benchmarks, and a PhD-level reasoning dataset. Compared to initial single-model attempts, ICE improved final overall accuracy by up to 27%. It reached accuracies 81% in medical subsets and 72% in multi-domain tasks from initial scores of about 72% and 60%, respectively. In a particularly challenging PhD-level reasoning benchmark (GPQA-diamond), ICE raised performance from 46.9% initially to 68.2% at the final consensus, a relative gain exceeding 45%. On a specialized family medicine dataset, ICE’s results were statistically indistinguishable from those of a complex reasoning model (O1-preview), despite O1’s higher cost and computational demands. Additional analyses showed that ICE’s iterative consensus remained effective under different prompting styles. Our proposed framework leverages standard LLMs and repeated prompting, requiring no specialized reward models or intricate token-level fusion. These findings show that iterative collaboration can transform LLM ensembles into more reliable, cost-efficient solvers, advancing performance in medical and general reasoning domains. Future refinements may integrate chain-of-thought steps or specialist models, extending this approach to more complex challenges as LLMs and benchmarks continue to evolve.

## Introduction

AI and large language models (LLMs) are showing competent performances in various domains (1–3). Benchmarks evaluating LLM performance in medical question answering, such as specialized multiple-choice exams (MCQs), reveal that some models now match or surpass human test-takers, and similar gains are seen in multi-domain question sets (4,5). Yet, while earlier transitions—such as from GPT-3.5 to GPT-4—yielded significant leaps, recent advancements, such as moving from GPT-4 to GPT-4o, have been more incremental (6–8). This reflects a broader trend: scaling models further does not always translate into proportionally better outcomes (9).

In sensitive areas like medicine, the stakes are high. Although LLMs achieve impressive accuracy on standardized tests, real-world applications demand even higher reliability. Persistent issues remain, including occasional hallucinations, flawed “reasoning” patterns, and inaccuracies that could misguide clinical decision-making (10,11). Thus, even as models demonstrate human-level performance on certain benchmarks, they may still falter under more complex, ambiguous, or nuanced conditions.

The current landscape also includes emerging “reasoning” models like openAI’s O1, Alibaba’s QWQ and Google’s Gemini 2.0 flash thinking (12,13). These often promise more structured thinking but come at the cost of longer processing times, greater computational demands, and higher expenses (12,13). They raise fundamental questions about whether models truly reason or merely produce convincing imitations of reasoning processes (14). The O1 model reportedly requires up to 10 times more compute during inference time, leading to significantly higher latency and costs (13).

Our proposed method, *Iterative Consensus Ensemble (ICE)*, diverges from this path. Rather than relying on a single model’s advancements, we combine multiple LLMs, guiding them through iterative explanation and refinement steps. This ensembling method is still somewhat resource-heavy, especially when state-of-the-art LLMs are involved. Yet, ongoing trends toward more efficient and faster models—smaller architectures becoming smarter (e.g., QWQ 32B-preview, LLaMA 3.3 70B)—could make such approaches increasingly feasible (15). If ICE proves effective, it may open new avenues for improving accuracy and consistency, reducing hallucinations, and enhancing real-world reliability through collective verification.

Various methods have explored LLM ensembling to improve reasoning and accuracy. Some approaches fuse probability distributions at each decoding step, converting different LLM outputs into a shared relative space to select improved tokens (16). Others treat ensembling as a form of weak-to-strong generalization, where weaker models trained on simpler tasks collectively guide a stronger model on harder questions (17). Another line of work frames the problem as a dynamic selection process, modeling ensemble as a sequence of decisions and using knowledge transfer prompts to pass information along chosen routes (18). Finally, another framework rely on pairwise ranking of candidate outputs and then merge the best solutions into a single, improved answer (19). We draw upon these ideas yet differ by employing an iterative, explanation-driven process that encourages diverse models to refine each other’s reasoning steps until they converge on a shared consensus.

We aim to assess whether an ensemble of LLMs, exchanging reasoning steps and converging on a consensus answer, can outperform individual models and improve upon their standalone iterations. In doing so, we seek to demonstrate that iterative collaboration among LLMs may be a viable strategy for pushing performance forward, even as direct leaps from one model generation to the next diminish.

## Materials and methods

### Overview of Study Design

We implemented an iterative ensemble method to improve the accuracy of LLMs on diverse multiple-choice question (MCQ) sets. We used four datasets (for a total of 4058 MCQs): (1) a newly curated set of 200 MCQs from the 2024 Israeli primary care licensing exam, referencing standard clinical sources (Goroll’s Primary Care Medicine and Nelson’s Textbook of Pediatrics) (20,21). It was created to ensure challenging content not widely available during model training; (2) a set of 655 open-access MCQs from Katz et al. (4), spanning internal medicine, general surgery, obstetrics and gynecology, pediatrics, and psychiatry; (3) a 3,000-question subset from the MMLU-PRO database, covering 12 distinct domains (biology, business, chemistry, computers, economics, history, law, math, philosophy, physics, engineering, and psychology – 250 random sample from each) (22); and (4) a collection of 198 PhD-level reasoning MCQs (GPQA-Diamond) commonly used to benchmark advanced reasoning models, serving as a validation set (23). All questions adhered to a single-best-answer format. Detailed dataset preparation and validation steps are described in the **supplementary materials**. **Figure 1** presents the study design and ICE framework.

**Figure 1.**
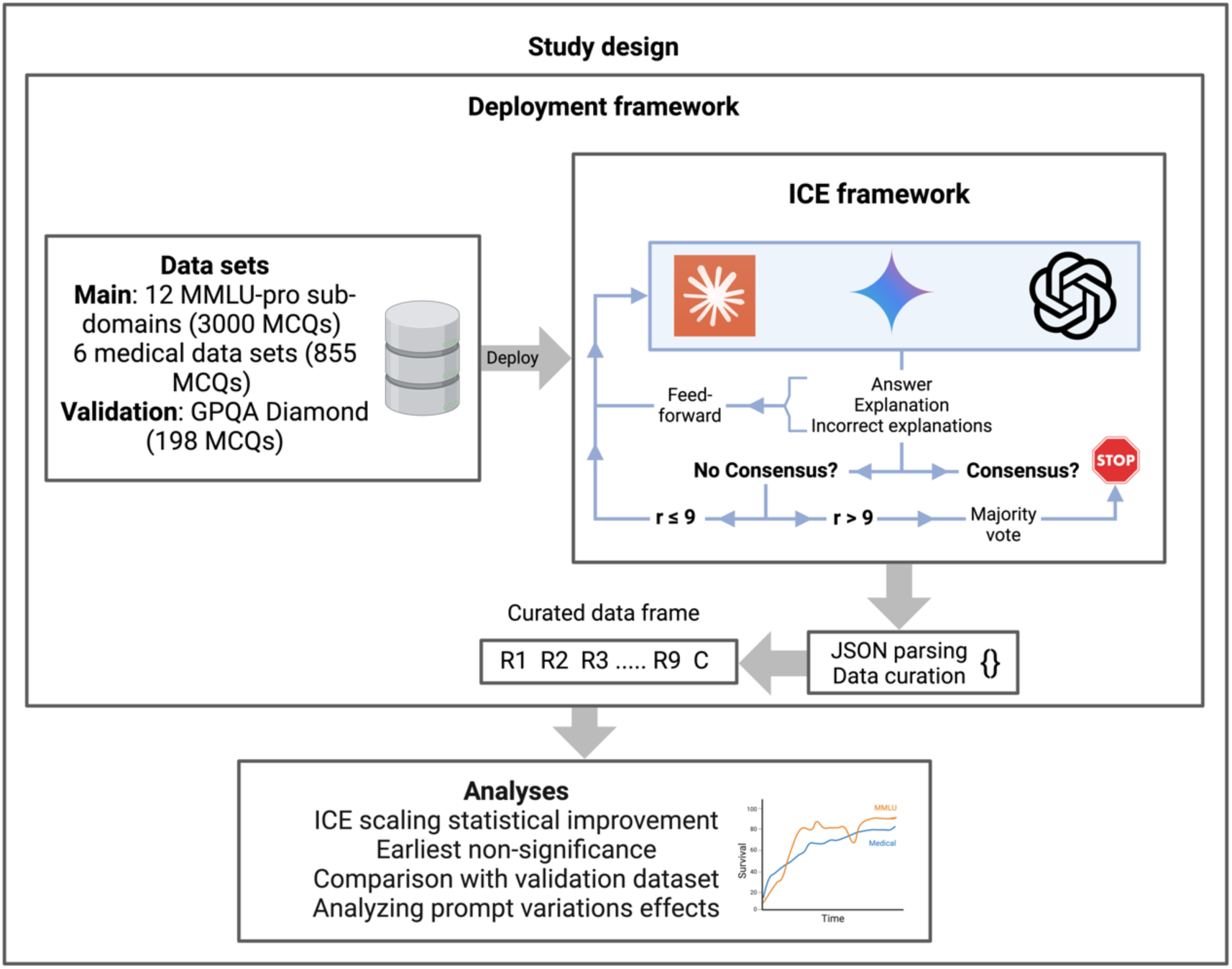
A flowchart of the study design.

### Model Selection and Ensemble Procedure

We selected three state-of-the-art LLMs: Claude Sonnet 3.5 (Anthropic), GPT-4o (OpenAI), and Gemini-1.5-pro (Google). Each model was first presented with a standardized prompt and was asked to produce an answer with reasoning, and justifications for excluding other options.

Each model was given a standardized prompt and produced an answer along with detailed reasoning for why it selected that answer and explanations for why other options were incorrect. The core principle of ICE is to iteratively refine each model’s answer by exposing it to the previous iteration’s reasoning and decisions of the other models, thereby guiding the three models toward a consensus.

We denote the three LLMs as m_1_, m_2_ and m_3_. Each question Q with answer set A = {a_1_, … , a_2_} underwent up to nine iterations:

Each model independently generates an initial answer: m_1,1_, m_2,1_ and m_3,1_. For iteration *r* (where 2 ≤ r ≤ 9), let:

- ξ_i,r_ denote the explanation from model *i* at iteration *r*.
- ω_i,r_ denote the incorrect-option explanations from model *i* at iteration *r*. The iterative process follows:

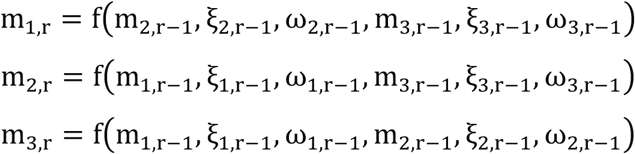

Where f(⋅) represents the model’s decision function based on other models’ previous outputs. The process terminates at iteration *r* if:

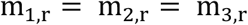

If no consensus emerges after nine iterations, we implement a majority voting system across all 27 decisions (3 models × 9 iterations). For each answer a ∈ A, we calculate:

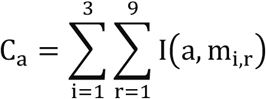

where the indicator function is defined as:

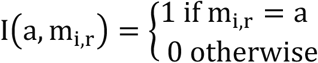

The final answer is determined by:

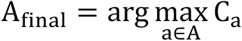

### Data Collection and Execution

We implemented the ICE approach in Python, using model-specific Application Programming Interfaces (APIs) and libraries. The code included JSON-based output validation, error handling, and retry logic. Each question was processed through up to three rounds of reasoning. The resulting answers, confidences, and reasoning were stored in Excel files for subsequent analysis.

Default hyper-parameters were used in all API calls. The entire code will be available and accessible on GitHub.

### Statistical Analysis

We computed mean accuracy values and 95% confidence intervals for each iteration and condition of interest. Normality of the paired differences was assessed using the Shapiro-Wilk test. If the data were normally distributed, we performed paired t-tests for pairwise comparisons. In the case of non-normal distributions, we employed the Wilcoxon signed-rank test. To control for multiple comparisons, p-values were adjusted using the Benjamini-Hochberg (BH) false discovery rate procedure, with a significance level of p < 0.05.

Additionally, we employed a mixed-effects logistic regression model (using question-level random intercepts and round as a fixed factor with round 9 as reference) to estimate differences in correctness at each round compared to the final (round 9) performance, identifying the earliest round at which no statistically significant deviation from final-round accuracy was observed. To determine the earliest iteration at which accuracy no longer differed significantly from the final consensus, we examined each round sequentially until no significant difference was observed. We also assessed model variability by tracking answer changes across rounds and tested differences in the frequencies of these transitions using analogous non-parametric comparisons when assumptions of normality were not met. All statistical analyses were conducted using R version 4.4.2.

### Stress testing and Validation in the Law Subset with Few-Shot Prompting and Chain of Thought (CoT)

As additional stress tests, we conducted a series of supplementary validation experiments on the law subset of the MMLU-Pro dataset. First, we introduced a few-shot prompt during the initial setup of the base models, thereby providing exemplar reasoning steps prior to any iterative refinement. In a second scenario, we applied a few-shot CoT style in the first run and enforced a strict CoT reasoning structure for each subsequent iteration, ensuring that all rounds followed a uniform and transparent explanation framework. In a third setup, we executed the ICE process using only the Claude model, running multiple iterations while the model remained unaware that it was interacting with its own previous responses. Finally, we repeated the standard ICE procedure on the same law subset to facilitate direct comparison. All these supplementary tests followed the same iterative evaluation protocol, statistical analysis methods, and data processing steps as our primary experiment.

## Results

### Overall Performance and Iterative Improvement

The Medical subset achieved a mean consensus score (the mean of the final answers of each ensemble run) of 81.17%, while MMLU reached 72.00%. The final consensus score for the entire dataset was 74.03%.

In round 1, the mean scores were 72.4% in Medical, 56.7% in MMLU, and 60.2% in the combined dataset. Compared to the initial scores, the consensus scores showed significant increases. Medical improved by 12.11% (absolute increase of 8.75 points, p < 0.001), MMLU by 27% (absolute 15.33 points, p < 0.001), and the combined dataset by 23% (absolute 13.84 points, p < 0.001) (**Figure 2**).

**Figure 2.**
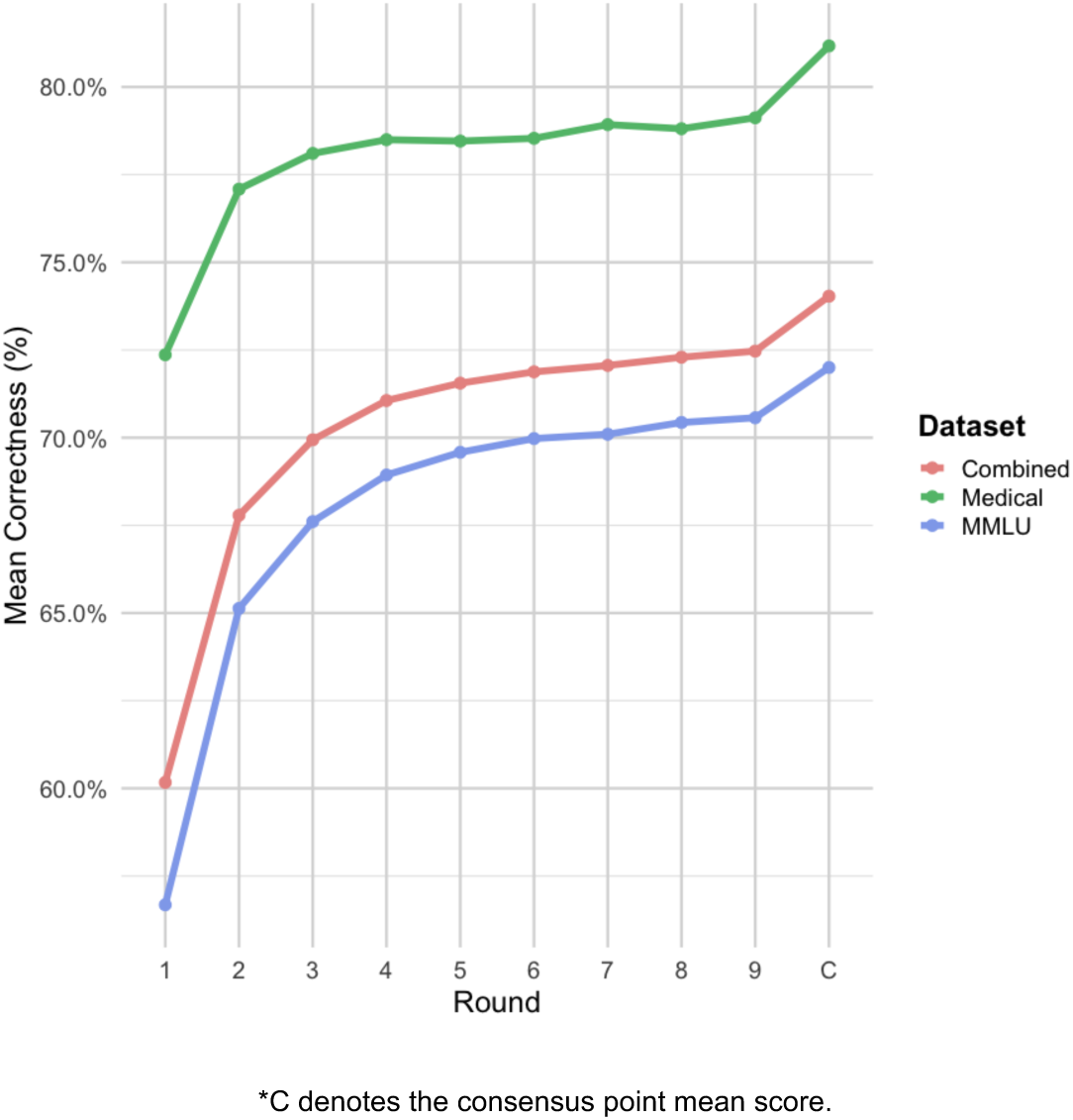
A summary of the mean performance across iterations and final weighted ensemble answer.

By round 5, the gaps between round 5 and consensus had decreased but remained significant. Medical was 2.61 points below consensus (p = 0.023), MMLU was 2.42 points short (p < 0.001), and the combined dataset trailed by 2.49 points (p < 0.001).

By round 9, Medical’s difference narrowed to 1.95 points (p = 0.1602, not significant), MMLU’s gap reduced to 1.44 points (p = 0.056), and the combined dataset retained a small but significant 1.58-point gap (p < 0.001).

From round 1 to round 9, the raw improvement in mean correctness was +6.7 percentage points in Medical (72.4% to 79.1%), +13.9 points in MMLU (56.7% to 70.6%), and +12.3 points in the combined dataset (60.2% to 72.5%). Regarding iterative progress, 45.32% of questions advanced beyond round 1, with fewer than 7% persisting to round 9. On average, questions concluded by round 2.29.

### Model-Specific Iteration Performance

Model-specific analyses indicated that all models started well below the consensus level and showed significant gains as rounds progressed, though these effects varied by dataset and model. For the Medical subset at round 1, Claude trailed consensus by 4.56 percentage points (p=0.0376), GPT-4o by 5.61 points (p=0.0093), and Gemini by 16.37 points (p<0.001). By round 5, none of these differences remained statistically significant except for Gemini’s marginal gap of 4.18 points (p=0.05), and by round 9, all three models approached consensus closely with no significant differences (**Table 1**).

**Table 1.** Model-Specific Iteration Comparisons (All, Medical, MMLU)

| Dataset | Model | R1 | R2 | R3 | R4 | R5 | R6 | R7 | R8 | R9 | C |
| --- | --- | --- | --- | --- | --- | --- | --- | --- | --- | --- | --- |
| Medical | Claude | 76.61 | 77.43 | 78.36 | 78.48 | 79.06 | 78.48 | 79.65 | 78.60 | 79.65 | 81.17 |
|  | GPT-4o | 75.56 | 76.73 | 79.77 | 78.95 | 79.30 | 78.71 | 79.65 | 79.30 | 79.18 |  |
|  | Gemini | 64.80 | 77.11 | 76.16 | 78.05 | 76.99 | 78.41 | 77.46 | 78.53 | 78.53 |  |
| MMLU | Claude | 56.99 | 66.12 | 67.66 | 69.39 | 69.52 | 69.96 | 70.09 | 70.36 | 70.89 | 72.00 |
|  | GPT-4o | 52.52 | 63.96 | 67.53 | 68.88 | 69.91 | 69.91 | 70.41 | 70.58 | 70.71 |  |
|  | Gemini | 60.62 | 65.31 | 67.63 | 68.52 | 69.31 | 70.05 | 69.78 | 70.36 | 70.09 |  |
| Combined | Claude | 61.34 | 68.63 | 70.03 | 71.41 | 71.64 | 71.85 | 72.21 | 72.18 | 72.83 | 74.03 |
|  | GPT-4o | 57.64 | 66.80 | 70.25 | 71.11 | 72.00 | 71.87 | 72.46 | 72.51 | 72.59 |  |
|  | Gemini | 61.56 | 67.94 | 69.53 | 70.65 | 71.02 | 71.92 | 71.49 | 72.18 | 71.97 |  |

\* R denotes round, and C denotes consensus.

In MMLU, round 1 differences were larger. Claude lagged by 15.01 points, GPT-4o by 19.48 points, and Gemini by 11.38 points (p<0.001 for all). By round 5, only Gemini remained significantly behind consensus, with a 2.69-point gap (p=0.0382), while Claude and GPT-4o were no longer significantly different. By round 9, no model significantly differed from consensus. In the combined data, all three models showed highly significant differences from consensus at round 1 (all p<0.001) and still had small but significant gaps by round 5 and round 9 (all p<0.001) (**Figure 3**).

**Figure 3.**
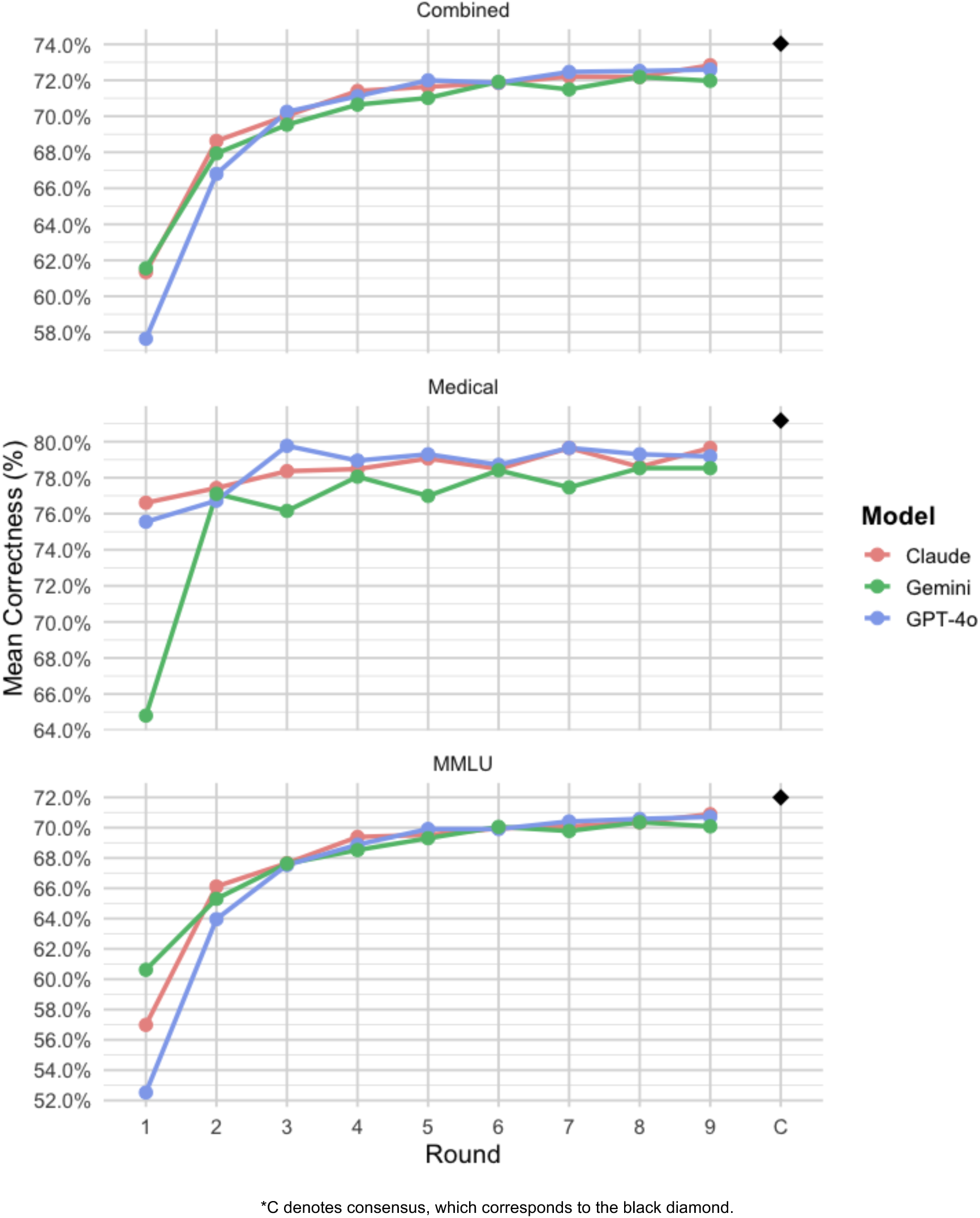
Model-specific overall performances by iteration.

### Validation via GPQA diamond dataset

In the validation dataset (GPQA-diamond), the consensus score averaged 68.18%. From an initial 46.9% at round 1 to 65.6% at round 9, the dataset showed a 39.9% improvement with 18.7 absolute point increase. Compared to the final consensus, round 1 was 21.04 points lower (p<0.001), round 5 still trailed by 5.39 points (p=0.0188), and by round 9 the difference narrowed to 2.53 points (p=0.257). These results reflect similar iterative improvement patterns observed in the primary datasets. (**Figure 3**).

**Figure 3.**
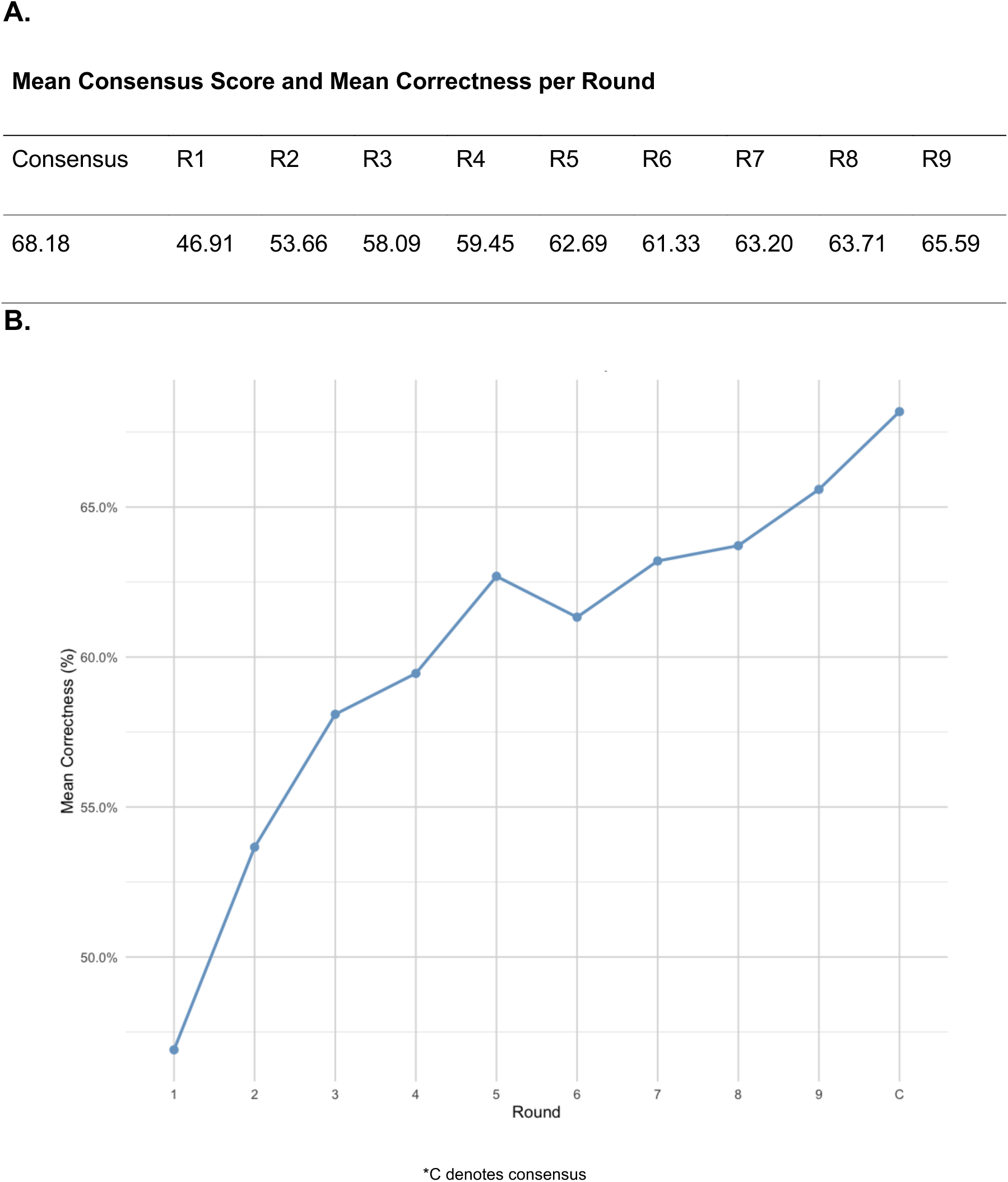
Overall iterated performance across rounds in the GPQA dataset. Prompt and model effects

### Stress testing and Validation in the Law Subset with Few-Shot Prompting and CoT

Additional validations using stress tests (as detailed in the methods section) in the law subset of MMLU, using a few-shot prompt for the base models (e.g. few-shot prompt was inputted to the models in the first round), achieved a consensus score of 63.6%, starting at 55.4% in round 1 and reaching 61.6% by round 9. Applying a few-shot CoT style in the first run and maintaining strict CoT explanations in subsequent iterations resulted in a 62.8% consensus, progressing from 55.5% at round 1 to 61.6% by round 9.

Running the ICE framework with Claude alone (“self-ensemble”) three times produced a 59.2% consensus, beginning at 58.5% and stabilizing around 59.3% in later rounds – lower than the score of the combined ensemble for this data base.

The standard ICE method on law data reached a 64.8% consensus, improving from 55.1% in round 1 to 63.6% by round 9. These additional results are generally consistent with the base ICE outcomes, showing no substantial differences in the observed iterative improvement patterns.

### Comparison of O1-preview and ICE Consensus in Family Medicine

Using our newly formatted and validated dataset of 200 family medicine MCQs, O1-preview-using the base prompt of the ICE framework-achieved a mean correctness of 82.5% [95% CI: 77.23%, 87.77%], while the ICE consensus reached 79.0% [95% CI: 73.35%, 84.65%] (**Figure 5**). Although O1’s performance was slightly higher, a Wilcoxon signed-rank test found no significant difference between the two approaches (p = 0.2269).

**Figure 4.**
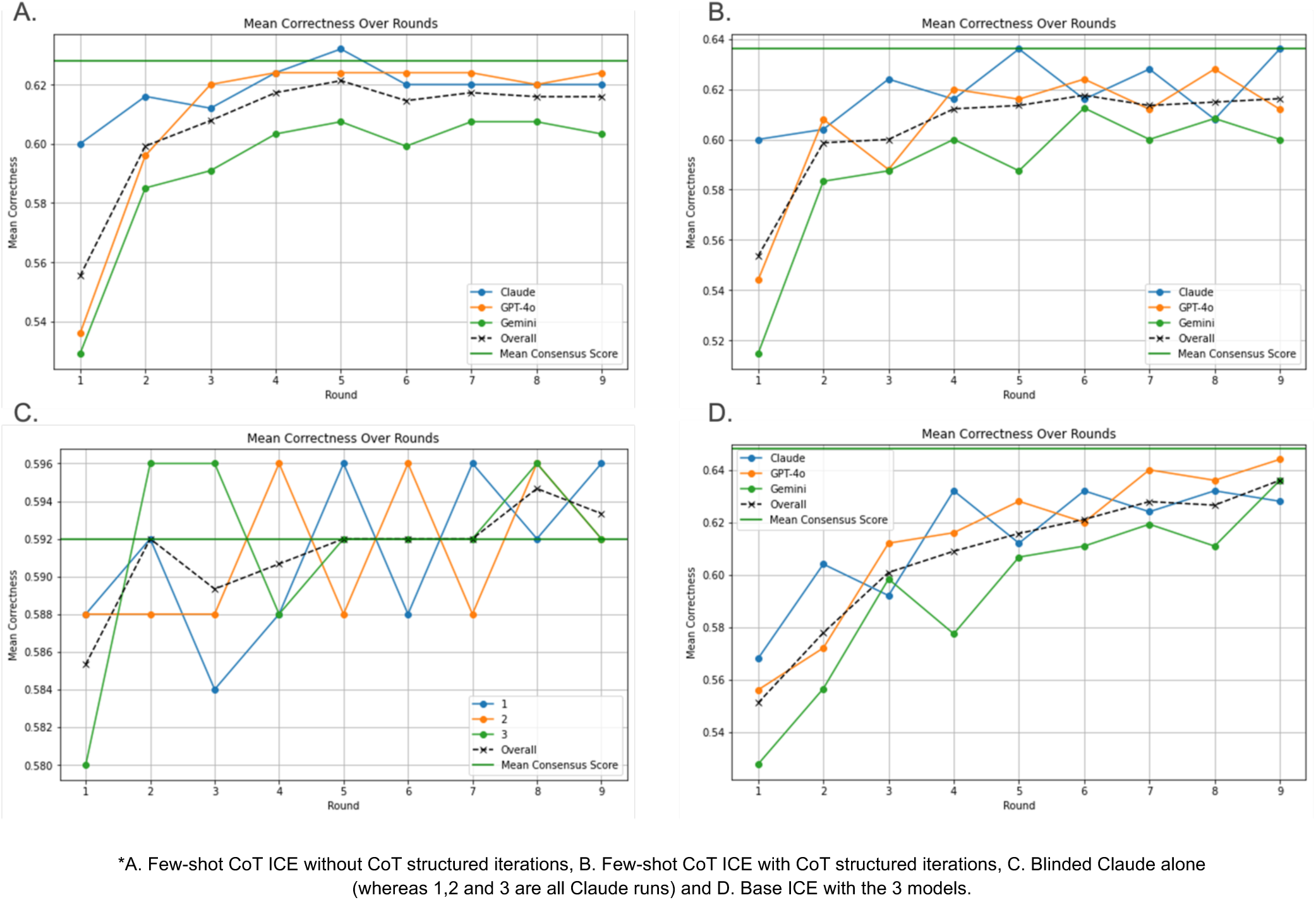
Results of the stress tests.

**Figure 5.**
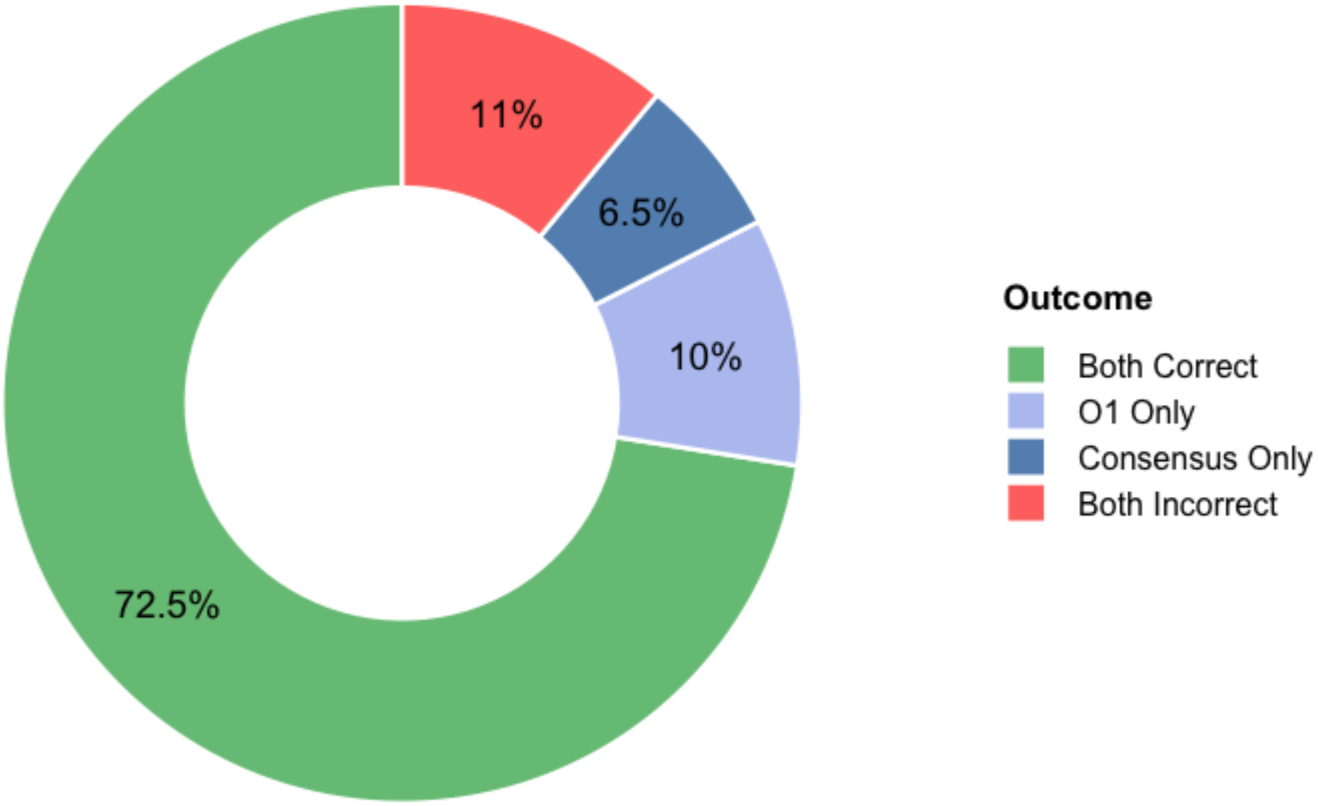
Question-specific performance comparison between O1-preview and ICE consensus scores.

## Discussion

The observed iterative improvements, seen over 30,000 prompted API runs, highlight the potential of guiding multiple LLMs toward a consensus answer through a structured, collaborative process. The entire dataset’s final consensus score reached 74.03%, reflecting a substantial-23%-gain from the initial combined accuracy of 60.2% at round 1. The Medical subset’s consensus climbed to 81.17% from an initial 72.4%, while MMLU rose to 72.00% from 56.7%. These gains were both statistically significant and meaningful in magnitude. This pattern of iterative refinement suggests that even well-performing models can benefit from additional rounds of reasoning and cross-evaluation. By round 5, the gap from consensus had narrowed but remained statistically significant for all datasets, indicating that a few extra iterations may still provide incremental improvements. By round 9, Medical no longer differed from its consensus score, MMLU’s difference was nearly negligible, and the combined dataset retained only a small, though statistically significant, advantage over its round 9 iteration. Most runs reached stable outcomes well before round 9, as on average, questions concluded by round 2.29, balancing the method’s iterative depth with practical efficiency. This is, to our knowledge, the largest attempt to ensemble LLMs for a medical reasoning benchmark, involving 855 questions from six different specialties, and including a newly curated, non-open-access dataset alongside other publicly available and validated sets. This approach may open the door to more refined, explainable, and accurate frameworks designed to make accessible healthcare through LLMs more feasible.

Importantly, model-specific analyses confirm that all chosen models— Claude Sonnet 3.5, GPT-4o, and Gemini 1.5 pro—started below the consensus level but approached it closely with only a handful of iterations. Although initial differences were larger in some subsets (e.g., MMLU) and varied across models, the iterative steps steadily reduced these gaps. By the later rounds, model-level differences from the consensus became modest or nonsignificant. Gemini, which often showed a larger initial gap, still aligned with the consensus after several refinement steps. This pattern suggests that even models starting from weaker initial positions can converge toward an improved standard through iterative reasoning. The results show that an ensemble approach, grounded in iterative exchange and refinement, can yield more stable and reliable answers, regardless of the models’ initial performance levels.

Additional stress tests on the law subset of MMLU-pro suggest that initial conditions and prompting strategies do not materially alter the iterative improvement patterns. Few-shot CoT prompting, whether applied initially or maintained throughout the iterations, elevated the starting accuracy but did not lead to different final outcomes. The method remained stable, indicating that ICE can operate effectively even with simple structured prompts.

Running ICE with only one model (Claude Sonnet 3.5), as a “self-ensemble” failed to replicate the multi-model gains. Although Claude started well, its performance plateaued at a lower consensus score and showed inconsistent shifts between rounds. This reinforces that ICE’s advantage arises from blending the diverse reasoning paths of multiple models, rather than relying on incremental refinements of a single LLM.

Our findings suggest that iterative consensus ensembling occupies a middle ground between simpler prompting strategies and more resource-intensive reasoning models. Prior works on CoT prompting and few-shot techniques highlight that providing richer context or carefully engineered prompts can help models reason more effectively (5,24,25). Still, these approaches can be sensitive to the selection and complexity of prompts, the number of examples, or the domain specificity of the instructions (24,26,27). In contrast, our framework seems less reliant on fine-tuned initial conditions. We see stable improvement patterns even when starting from relatively generic prompts, implying that the iterative feedback from multiple models may mitigate the delicate prompt variations known to influence CoT outcomes.

Other ensemble methods have explored complex token-level distributions or extra reward models to fuse diverse outputs from multiple LLMs (16,19). While these methods yield gains, their dependency on intricate calibration and alignment procedures may limit their adaptability across new tasks or unseen data distributions. Our simpler, round-by-round comparison and refinement strategy offers a more direct route to leveraging complementary model strengths. In a related vein, Barabucci et al. tested 200 real-life clinical vignettes from the Human Diagnosis Project and aggregated differential diagnoses from multiple commercial LLMs (e.g., GPT-4, PaLM 2, Llama 2, Cohere Command) using a frequency-based weighting method that assigned more weight to repeatedly appearing or top-ranked answers. This raised top-5 diagnostic accuracy from an average 59.0% (single LLM) to 75.3% (ensemble). They also found that smaller, less capable LLM combinations approached GPT-4’s performance, reducing reliance on any single vendor (28). Similarly, dynamic ensemble reasoning and adaptive selection methods, as suggested by Hu et al., often involve sophisticated decision structures or reinforcement learning to choose the best candidate model at runtime (18). Our approach differs by focusing on emergent consensus through iterative negotiation, reducing the need for model-specific heuristics or extensive domain engineering.

Comparing our approach to specialized large “inference-time compute” models (e.g., O1-preview) shows dataset-dependent patterns. On our newly introduced family medicine benchmark of previously unavailable MCQs, the O1-preview and ICE consensus scores are statistically indistinguishable. However, on the GPQA diamond dataset, our final accuracy (68%) appears to fall slightly behind O1-preview’s reported ∼74%, though the lack of direct statistical testing with their reported results makes firm conclusions difficult. This suggests that while our iterative ensemble can match O1-preview in certain specialized medical domains, its relative standing may vary across other contexts. Yet, the resource demands for O1-preview—both in terms of compute time and cost— remain substantially greater than those required by our proposed ICE framework. O1-preview, which is tailored for multi-step reasoning and typically generates longer chains of thought, can cost up to six times more per token compared to standard GPT-4o, and in some evaluations produces more than twice the output tokens for similar tasks (29). Moreover, global cost assessments from other studies indicate that O1-preview might be 65 times more expensive than GPT-4o and up to 95 times more than Sonnet 3.5 performing the same “reasoning” tasks (29). By comparison, our iterative ensemble framework relies on accessible models and repeated calls without a specialized reasoning backend, potentially offering a more cost-effective means of approaching advanced reasoning capabilities. Our proposed framework does not fully match the strongest reasoning models on every metric, but we may narrow the gap at a fraction of the price and complexity. Future refinements might further integrate CoT steps, domain-specific heuristics, or specialist models into our ensemble, enhancing performance without incurring the prohibitive costs and complexities associated with current large reasoning models. Especially with fast evolving reasoning models, that are smaller in scale, more efficient and produce consistent, and sometimes even better, results in benchmarks compare to O1 and other state-of-the-art larger models, like QwQ-32B (12).

Our study has limitations worth acknowledging. First, the ICE framework relies on multiple proprietary models, leaving open questions about its efficacy when applied to smaller, open-source LLMs or domain-specific reasoning models. Exploring ensembles composed of lighter, locally hosted models could broaden accessibility and test whether performance gains persist under more constrained computational or licensing conditions. Second, our consensus strategy currently checks for agreement at fixed iteration endpoints, often at round one; adjusting this trigger point—such as waiting until round two or three before finalizing consensus—might yield smoother or more stable improvement curves. Third, we have focused on a subset of tasks and domains, including medical and multi-domain MCQs, while leaving unexplored a wide range of reasoning benchmarks, coding challenges, and emergent creative tasks that could further illuminate the method’s strengths and weaknesses. Finally, as reasoning models evolve, the benchmarks must follow suit (30). Developing more demanding, nuanced, and standardized evaluations will help clarify whether iterative ensembling consistently adds value and how best to fine-tune it.

In conclusion, our proof-of-concept demonstrates that an iterative, consensus-driven ensemble of multiple LLMs can enhance accuracy beyond what individual models achieve on their own, even matching specialized large reasoning models like O1-preivew. By letting models refine each other’s reasoning in successive rounds, the ensemble converges toward a stable consensus that significantly outperforms initial standalone attempts – up to more than 20%. This improvement persists across multiple datasets and arises even when starting from simpler prompts. Taken together, these findings suggest that iterative ensembling may be a practical, adaptable pathway for pushing medical and general LLMs tasks toward more reliable, consistent answers.

## Data Availability

All data produced in the present study are available upon reasonable request to the authors

## Ethical approval

was not required for this research as only synthetic open-access data was used.

## Contributorship Statement

MO led the study design, data analysis, visualizations, and manuscript drafting. BSG, GNN, and EK all contributed significantly to the project, editing and revising the manuscript.

