## Supplementary Materials for "Refining LLMs Outputs with Iterative Consensus Ensemble (ICE)"

| Data Set | Model | Comparison | Mean Difference | Adjusted p value |
|---|---|---|---|---|
| **All Datasets** | GPT-4o | Iteration 1 vs Iteration 2 | 10.6% | <0.001 |
| **All Datasets** | GPT-4o | Iteration 1 vs Iteration 3 | 12.7% | <0.001 |
| **All Datasets** | GPT-4o | Iteration 2 vs Iteration 3 | 2.1% | <0.001 |
| **All Datasets** | GPT-4o | Final vs Iteration 1 | 13.1% | <0.001 |
| **All Datasets** | GPT-4o | Final vs Iteration 2 | 2.5% | <0.001 |
| **All Datasets** | GPT-4o | Final vs Iteration 3 | 0.3% | <0.001 |
| **All Medical** | GPT-4o | Iteration 1 vs Iteration 2 | 3.2% | <0.001 |
| **All Medical** | GPT-4o | Iteration 1 vs Iteration 3 | 5.3% | <0.001 |
| **All Medical** | GPT-4o | Iteration 2 vs Iteration 3 | 2.0% | <0.001 |
| **All Medical** | GPT-4o | Final vs Iteration 1 | 6.1% | <0.001 |
| **All Medical** | GPT-4o | Final vs Iteration 2 | 2.9% | <0.001 |
| **All Medical** | GPT-4o | Final vs Iteration 3 | 0.8% | 0.04 |
| **All MMLU** | GPT-4o | Iteration 1 vs Iteration 2 | 13.2% | <0.001 |
| **All MMLU** | GPT-4o | Iteration 1 vs Iteration 3 | 15.4% | <0.001 |
| **All MMLU** | GPT-4o | Iteration 2 vs Iteration 3 | 2.2% | <0.001 |
| **All MMLU** | GPT-4o | Final vs Iteration 1 | 15.5% | <0.001 |
| **All MMLU** | GPT-4o | Final vs Iteration 2 | 2.3% | 0.003 |
| **All MMLU** | GPT-4o | Final vs Iteration 3 | 0.2% | 0.071 |
| **All Datasets** | Claude | Iteration 1 vs Iteration 2 | 10.3% | <0.001 |
| **All Datasets** | Claude | Iteration 1 vs Iteration 3 | 13.3% | <0.001 |
| **All Datasets** | Claude | Iteration 2 vs Iteration 3 | 3.0% | <0.001 |
| **All Datasets** | Claude | Final vs Iteration 1 | 15.5% | <0.001 |

| | | | | |
|---|---|---|---|---|
| **All Datasets** | Claude | Final vs Iteration 2 | 5.2% | <0.001 |
| **All Datasets** | Claude | Final vs Iteration 3 | 2.2% | <0.001 |
| **All Medical** | Claude | Iteration 1 vs Iteration 2 | 3.4% | <0.001 |
| **All Medical** | Claude | Iteration 1 vs Iteration 3 | 4.3% | <0.001 |
| **All Medical** | Claude | Iteration 2 vs Iteration 3 | 1.0% | <0.001 |
| **All Medical** | Claude | Final vs Iteration 1 | 6.5% | <0.001 |
| **All Medical** | Claude | Final vs Iteration 2 | 3.1% | <0.001 |
| **All Medical** | Claude | Final vs Iteration 3 | 2.2% | 0.012 |
| **All MMLU** | Claude | Iteration 1 vs Iteration 2 | 12.7% | <0.001 |
| **All MMLU** | Claude | Iteration 1 vs Iteration 3 | 16.5% | <0.001 |
| **All MMLU** | Claude | Iteration 2 vs Iteration 3 | 3.7% | <0.001 |
| **All MMLU** | Claude | Final vs Iteration 1 | 18.7% | <0.001 |
| **All MMLU** | Claude | Final vs Iteration 2 | 6.0% | <0.001 |
| **All MMLU** | Claude | Final vs Iteration 3 | 2.3% | 0.015 |
| **All Datasets** | Gemini | Iteration 1 vs Iteration 2 | 2.4% | <0.001 |
| **All Datasets** | Gemini | Iteration 1 vs Iteration 3 | 4.2% | <0.001 |
| **All Datasets** | Gemini | Iteration 2 vs Iteration 3 | 1.7% | <0.001 |
| **All Datasets** | Gemini | Final vs Iteration 1 | 8.2% | <0.001 |
| **All Datasets** | Gemini | Final vs Iteration 2 | 5.8% | <0.001 |
| **All Datasets** | Gemini | Final vs Iteration 3 | 4.1% | <0.001 |
| **All Medical** | Gemini | Iteration 1 vs Iteration 2 | 9.7% | <0.001 |
| **All Medical** | Gemini | Iteration 1 vs Iteration 3 | 11.0% | <0.001 |
| **All Medical** | Gemini | Iteration 2 vs Iteration 3 | 1.3% | <0.001 |
| **All Medical** | Gemini | Final vs Iteration 1 | 15.0% | <0.001 |
| **All Medical** | Gemini | Final vs Iteration 2 | 5.3% | <0.001 |

| | | | | |
|---|---|---|---|---|
| **All Medical** | Gemini | Final vs Iteration 3 | 4.0% | <0.001 |
| **All MMLU** | Gemini | Iteration 1 vs Iteration 2 | -0.1% | 0.813 |
| **All MMLU** | Gemini | Iteration 1 vs Iteration 3 | 1.8% | <0.001 |
| **All MMLU** | Gemini | Iteration 2 vs Iteration 3 | 1.9% | <0.001 |
| **All MMLU** | Gemini | Final vs Iteration 1 | 5.9% | <0.001 |
| **All MMLU** | Gemini | Final vs Iteration 2 | 6.0% | <0.001 |
| **All MMLU** | Gemini | Final vs Iteration 3 | 4.1% | <0.001 |

### Statistical Determination of the Earliest Round Achieving Performance Stabilization

#### Methodological Overview

*Data Structure and Setting:* Our objective is to determine the earliest round at which the ensemble's correctness is statistically equivalent to that at Round 9, hence indicating no further significant gains in performance.

*Outcome Variable:* The outcome of interest is binary correctness (0/1) for each question at each round. Correctness is defined as whether the chosen ensemble answer matches the ground-truth answer. Thus, each question contributes up to nine repeated binary outcomes (one per round), with Round 9 considered the benchmark or "final" level of performance.

#### Statistical Model

*Mixed-Effects Logistic Regression:* We modeled correctness as a function of the round number using a generalized linear mixed model (GLMM) with a binomial (logistic) link. In formula terms:

$$\mathrm{Correctness}_{ij} \sim \mathrm{Bernoulli}(p_{ij}), \mathrm{logit}(p_{ij}) = \beta_0 + \beta_{\mathrm{Round}=r} + u_{q_i}$$

where $p_{i,j}$ is the probability of correctness for question I at round j, $\beta_0$ is the baseline log-odds of correctness at the reference round, and $\beta_{\mathrm{round}=r}$ represents the fixed effect of the round r (compared to the reference round). We included a random intercept $u_{q_i}$ for each question i to account for question-level variability and repeated measures.

*Choice of Reference Category:* We designated Round 9 as the reference round. This allows all pairwise comparisons to be interpreted as differences relative to the final round performance. Consequently, each earlier round's performance is tested against the Round 9 benchmark.

*Estimation and Software:* We used R's lme4 package (main function: glmer()) to fit the logistic GLMM. This approach efficiently estimates fixed effects and random intercepts, handling repeated measures and accounting for the clustered structure of data at the question level.

#### Post-Hoc Comparisons and Significance Testing

*Marginal Means and Contrasts:* After fitting the GLMM, we estimated the marginal means of correctness for each round using the emmeans package (main functions: emmeans() and contrast()). Marginal means represent the model-adjusted probability of correctness at each round, averaged over the random effects.

*Multiple Comparisons and Adjustment Method:* To determine the earliest round that does not significantly differ from Round 9, we performed pairwise comparisons between each earlier round and Round 9 using contrast(..., method="revpairwise"). This approach generates comparisons of each round with the reference round. We applied Tukey's adjustment for multiple comparisons. Tukey's method is well-established for controlling the family-wise error rate when performing pairwise comparisons among multiple groups. By using a Tukey correction, we ensure that the reported p-values for comparisons remain valid after accounting for the multiple testing scenario (eight comparisons in this case, one for each earlier round versus Round 9).

*Interpretation of Non-Significance:* If a given round does not differ significantly from Round 9 (p-value > 0.05, after Tukey adjustment), we conclude that the ensemble performance at that round is statistically indistinguishable from the final consensus performance. By identifying the earliest such round, we pinpoint where no further statistically meaningful improvement occurs, providing a practical stopping point for the iterative process.

**Identification of the Earliest Non-Significant Round**

*Procedure:*

1. Fit the mixed-effects logistic model using glmer().
2. Obtain marginal means for each round using emmeans().
3. Compute pairwise comparisons of each earlier round to Round 9 via contrast().
4. Apply Tukey-adjusted p-values to control for multiple comparisons.
5. Identify all rounds that yield non-significant differences from Round 9.
6. Select the earliest (lowest-numbered) round from this set as the earliest non-significant round.

If no earlier rounds are non-significant, it implies that performance continually improves (or is at least statistically distinguishable) until the final round.

**Summarizing Results**

The tables of raw contrasts (estimates, standard errors, z-ratios, and Tukey-adjusted p-values) comparing each round's performance to the final reference round (Round 9) revealed distinct patterns of when performance ceased to differ significantly from that terminal benchmark.

In the medical dataset, the logistic mixed model estimated that, while Rounds 1 and 2 were significantly lower than Round 9 ($p < 0.0001$ in both cases), by Round 3 the difference became non-significant ($p \approx 0.9036$ when compared to Round 2, which had already established proximity to Round 9). In other words, Round 3 demonstrated no statistically meaningful performance gap relative to Round 9. Continuing through Rounds 4, 5, and beyond, the tests also indicated that there were no newly significant divergences once stability had been reached. Thus, the medical dataset's performance stabilized at Round 3.

For the MMLU dataset, the process of stabilization required more rounds. While early rounds (e.g., Round 2 vs. Round 9) still displayed strong statistical differences ($p < 0.0001$), the earliest round to show a non-significant difference relative to Round 9 emerged only at Round 5 ($p \approx 0.5149$ when compared to Round 4, which was already close to Round 9's level of performance). Similar subsequent comparisons (e.g., Round 6, Round 7, and so forth) provided further confirmation that once Round 5 was reached, no further significant improvements occurred. Therefore, the MMLU dataset stabilized at Round 5.

A similar pattern emerged in the combined dataset analysis. After prominent differences in earlier rounds, Round 5 was the first point at which no statistically significant difference from Round 9 persisted ($p \approx 0.1777$ for the direct comparison of Round 5 vs. Round 9). Examination of subsequent rounds indicated that no additional significant improvements or declines materialized. Consequently, the combined dataset achieved stabilization by Round 5.

Quantitatively, the pattern was clear: earlier rounds displayed strongly significant differences from the final benchmark (e.g., Round 1 vs. Round 9 often with $p < 0.0001$ and large negative log-odds estimates), while as we progressed to Rounds 3–5, the contrasts diminished in magnitude and eventually lost statistical significance.

Table S*: Medical - Raw Contrast Results.

| Contrast | Tested Round | Estimate (log-odds diff) | Std. Error | Z-Ratio | P-Value |
|---|---|---|---|---|---|
| Round1 - Round9 | 1 | −2.632 | 0.166 | −15.876 | 0.0000 |
| Round2 - Round9 | 2 | −1.331 | 0.168 | −7.913 | 0.0000 |
| Round2 - Round1 | 2 | 1.302 | 0.148 | 8.798 | 0.0000 |
| Round3 - Round9 | 3 | −1.114 | 0.169 | −6.590 | 0.0000 |
| Round3 - Round1 | 3 | 1.519 | 0.151 | 10.080 | 0.0000 |
| Round3 - Round2 | 3 | 0.217 | 0.157 | 1.386 | 0.9036 |
| Round4 - Round9 | 4 | −0.839 | 0.170 | −4.931 | 0.0000 |

| Contrast | Tested Round | Estimate (log-odds diff) | Std. Error | Z-Ratio | P-Value |
|---|---|---|---|---|---|
| Round4 - Round1 | 4 | 1.794 | 0.154 | 11.633 | 0.0000 |
| Round4 - Round2 | 4 | 0.492 | 0.159 | 3.092 | 0.0515 |
| Round4 - Round3 | 4 | 0.275 | 0.161 | 1.713 | 0.7390 |
| Round5 - Round9 | 5 | −0.776 | 0.170 | −4.550 | 0.0002 |
| Round5 - Round1 | 5 | 1.857 | 0.155 | 11.977 | 0.0000 |
| Round5 - Round2 | 5 | 0.555 | 0.160 | 3.475 | 0.0150 |
| Round5 - Round3 | 5 | 0.338 | 0.161 | 2.099 | 0.4742 |
| Round5 - Round4 | 5 | 0.063 | 0.163 | 0.387 | 1.0000 |
| Round6 - Round9 | 6 | −0.965 | 0.170 | −5.692 | 0.0000 |
| Round6 - Round1 | 6 | 1.667 | 0.153 | 10.927 | 0.0000 |
| Round6 - Round2 | 6 | 0.365 | 0.158 | 2.313 | 0.3341 |
| Round6 - Round3 | 6 | 0.148 | 0.159 | 0.930 | 0.9913 |
| Round6 - Round4 | 6 | −0.127 | 0.162 | −0.784 | 0.9973 |
| Round6 - Round5 | 6 | −0.190 | 0.162 | −1.171 | 0.9627 |
| Round7 - Round9 | 7 | −0.773 | 0.170 | −4.534 | 0.0002 |
| Round7 - Round1 | 7 | 1.860 | 0.155 | 11.992 | 0.0000 |
| Round7 - Round2 | 7 | 0.558 | 0.160 | 3.491 | 0.0142 |
| Round7 - Round3 | 7 | 0.341 | 0.161 | 2.115 | 0.4632 |
| Round7 - Round4 | 7 | 0.066 | 0.163 | 0.403 | 1.0000 |

| Contrast | Tested Round | Estimate (log-odds diff) | Std. Error | Z-Ratio | P-Value |
|---|---|---|---|---|---|
| Round7 - Round5 | 7 | 0.003 | 0.164 | 0.016 | 1.0000 |
| Round7 - Round6 | 7 | 0.192 | 0.162 | 1.187 | 0.9595 |
| Round8 - Round9 | 8 | −0.710 | 0.171 | −4.163 | 0.0010 |
| Round8 - Round1 | 8 | 1.922 | 0.156 | 12.330 | 0.0000 |
| Round8 - Round2 | 8 | 0.620 | 0.160 | 3.868 | 0.0035 |
| Round8 - Round3 | 8 | 0.403 | 0.162 | 2.494 | 0.2347 |
| Round8 - Round4 | 8 | 0.128 | 0.164 | 0.784 | 0.9973 |
| Round8 - Round5 | 8 | 0.065 | 0.164 | 0.397 | 1.0000 |
| Round8 - Round6 | 8 | 0.255 | 0.163 | 1.567 | 0.8229 |
| Round8 - Round7 | 8 | 0.062 | 0.164 | 0.381 | 1.0000 |
| **Earliest Non-Significant Round: 3** | **3** | **NA** | **NA** | **NA** | **NA** |

Table S*: MMLU - Raw Contrast Results

| Contrast | Tested Round | Estimate (log-odds diff) | Std. Error | Z-Ratio | P-Value |
|---|---|---|---|---|---|
| Round1 - Round9 | 1 | −3.221 | 0.080 | −40.173 | 0.0000 |
| Round2 - Round9 | 2 | −1.571 | 0.080 | −19.637 | 0.0000 |
| Round2 - Round1 | 2 | 1.650 | 0.069 | 24.074 | 0.0000 |
| Round3 - Round9 | 3 | −0.965 | 0.082 | −11.838 | 0.0000 |
| Round3 - Round1 | 3 | 2.256 | 0.073 | 30.956 | 0.0000 |
| Round3 - Round2 | 3 | 0.606 | 0.074 | 8.179 | 0.0000 |
| Round4 - Round9 | 4 | −0.587 | 0.083 | −7.106 | 0.0000 |
| Round4 - Round1 | 4 | 2.634 | 0.076 | 34.762 | 0.0000 |
| Round4 - Round2 | 4 | 0.984 | 0.076 | 12.884 | 0.0000 |
| Round4 - Round3 | 4 | 0.378 | 0.078 | 4.823 | 0.0000 |
| Round5 - Round9 | 5 | −0.422 | 0.083 | −5.062 | 0.0000 |
| Round5 - Round1 | 5 | 2.799 | 0.077 | 36.274 | 0.0000 |
| Round5 - Round2 | 5 | 1.149 | 0.078 | 14.823 | 0.0000 |
| Round5 - Round3 | 5 | 0.543 | 0.079 | 6.841 | 0.0000 |
| Round5 - Round4 | 5 | 0.165 | 0.081 | 2.040 | 0.5149 |
| Round6 - Round9 | 6 | −0.296 | 0.084 | −3.530 | 0.0124 |

| Contrast | Tested Round | Estimate (log-odds diff) | Std. Error | Z-Ratio | P-Value |
|---|---|---|---|---|---|
| Round6 - Round1 | 6 | 2.925 | 0.078 | 37.401 | 0.0000 |
| Round6 - Round2 | 6 | 1.275 | 0.078 | 16.262 | 0.0000 |
| Round6 - Round3 | 6 | 0.669 | 0.080 | 8.350 | 0.0000 |
| Round6 - Round4 | 6 | 0.291 | 0.082 | 3.569 | 0.0108 |
| Round6 - Round5 | 6 | 0.126 | 0.082 | 1.531 | 0.8412 |
| Round7 - Round9 | 7 | −0.249 | 0.084 | −2.963 | 0.0745 |
| Round7 - Round1 | 7 | 2.972 | 0.079 | 37.819 | 0.0000 |
| Round7 - Round2 | 7 | 1.322 | 0.079 | 16.800 | 0.0000 |
| Round7 - Round3 | 7 | 0.716 | 0.080 | 8.912 | 0.0000 |
| Round7 - Round4 | 7 | 0.338 | 0.082 | 4.138 | 0.0012 |
| Round7 - Round5 | 7 | 0.173 | 0.083 | 2.099 | 0.4737 |
| Round7 - Round6 | 7 | 0.047 | 0.083 | 0.569 | 0.9997 |
| Round8 - Round9 | 8 | −0.136 | 0.085 | −1.606 | 0.8019 |
| Round8 - Round1 | 8 | 3.086 | 0.080 | 38.773 | 0.0000 |
| Round8 - Round2 | 8 | 1.435 | 0.080 | 18.038 | 0.0000 |
| Round8 - Round3 | 8 | 0.829 | 0.081 | 10.218 | 0.0000 |

| Contrast | Tested Round | Estimate (log-odds diff) | Std. Error | Z-Ratio | P-Value |
|---|---|---|---|---|---|
| Round8 - Round4 | 8 | 0.451 | 0.082 | 5.475 | 0.0000 |
| Round8 - Round5 | 8 | 0.286 | 0.083 | 3.443 | 0.0168 |
| Round8 - Round6 | 8 | 0.160 | 0.084 | 1.916 | 0.6027 |
| Round8 - Round7 | 8 | 0.113 | 0.084 | 1.348 | 0.9167 |
| **Earliest Non-Significant Round: 5** | **5** | **NA** | **NA** | **NA** | **NA** |

Table S*: Combined - Raw Contrast Results.

| Contrast | Tested Round | Estimate (log-odds diff) | Std. Error | Z-Ratio | P-Value |
|---|---|---|---|---|---|
| Round1 - Round9 | 1 | −1.579 | 0.048 | −32.628 | 0.0000 |
| Round2 - Round9 | 2 | −0.629 | 0.048 | −13.071 | 0.0000 |
| Round2 - Round1 | 2 | 0.951 | 0.047 | 20.232 | 0.0000 |
| Round3 - Round9 | 3 | −0.348 | 0.049 | −7.169 | 0.0000 |
| Round3 - Round1 | 3 | 1.231 | 0.048 | 25.799 | 0.0000 |
| Round3 - Round2 | 3 | 0.281 | 0.048 | 5.885 | 0.0000 |
| Round4 - Round9 | 4 | −0.197 | 0.049 | −4.024 | 0.0019 |
| Round4 - Round1 | 4 | 1.383 | 0.048 | 28.718 | 0.0000 |
| Round4 - Round2 | 4 | 0.432 | 0.048 | 9.005 | 0.0000 |
| Round4 - Round3 | 4 | 0.152 | 0.048 | 3.129 | 0.0461 |
| Round5 - Round9 | 5 | −0.128 | 0.049 | −2.621 | 0.1777 |
| Round5 - Round1 | 5 | 1.451 | 0.048 | 30.056 | 0.0000 |
| Round5 - Round2 | 5 | 0.501 | 0.048 | 10.412 | 0.0000 |
| Round5 - Round3 | 5 | 0.220 | 0.049 | 4.536 | 0.0002 |
| Round5 - Round4 | 5 | 0.069 | 0.049 | 1.408 | 0.8953 |
| Round6 - Round9 | 6 | −0.083 | 0.049 | −1.696 | 0.7494 |
| Round6 - Round1 | 6 | 1.496 | 0.048 | 30.861 | 0.0000 |
| Round6 - Round2 | 6 | 0.546 | 0.048 | 11.303 | 0.0000 |

| Contrast | Tested Round | Estimate (log-odds diff) | Std. Error | Z-Ratio | P-Value |
|---|---|---|---|---|---|
| Round6 - Round3 | 6 | 0.265 | 0.049 | 5.437 | 0.0000 |
| Round6 - Round4 | 6 | 0.113 | 0.049 | 2.316 | 0.3324 |
| Round6 - Round5 | 6 | 0.045 | 0.049 | 0.911 | 0.9924 |
| Round7 - Round9 | 7 | −0.059 | 0.049 | −1.199 | 0.9570 |
| Round7 - Round1 | 7 | 1.521 | 0.049 | 31.309 | 0.0000 |
| Round7 - Round2 | 7 | 0.570 | 0.048 | 11.786 | 0.0000 |
| Round7 - Round3 | 7 | 0.290 | 0.049 | 5.937 | 0.0000 |
| Round7 - Round4 | 7 | 0.138 | 0.049 | 2.810 | 0.1123 |
| Round7 - Round5 | 7 | 0.069 | 0.049 | 1.407 | 0.8954 |
| Round7 - Round6 | 7 | 0.024 | 0.049 | 0.497 | 0.9999 |
| Round8 - Round9 | 8 | −0.025 | 0.049 | −0.507 | 0.9999 |
| Round8 - Round1 | 8 | 1.554 | 0.049 | 31.901 | 0.0000 |
| Round8 - Round2 | 8 | 0.604 | 0.048 | 12.459 | 0.0000 |
| Round8 - Round3 | 8 | 0.323 | 0.049 | 6.615 | 0.0000 |
| Round8 - Round4 | 8 | 0.172 | 0.049 | 3.494 | 0.0141 |
| Round8 - Round5 | 8 | 0.103 | 0.049 | 2.093 | 0.4780 |
| Round8 - Round6 | 8 | 0.058 | 0.049 | 1.182 | 0.9604 |
| Round8 - Round7 | 8 | 0.034 | 0.050 | 0.685 | 0.9990 |

| Contrast | Tested Round | Estimate (log-odds diff) | Std. Error | Z-Ratio | P-Value |
|---|---|---|---|---|---|
| **Earliest Non-Significant Round: 5** | **5** | **NA** | **NA** | **NA** | **NA** |

Figure S*: Correctness per round comparison across data sets and overall.

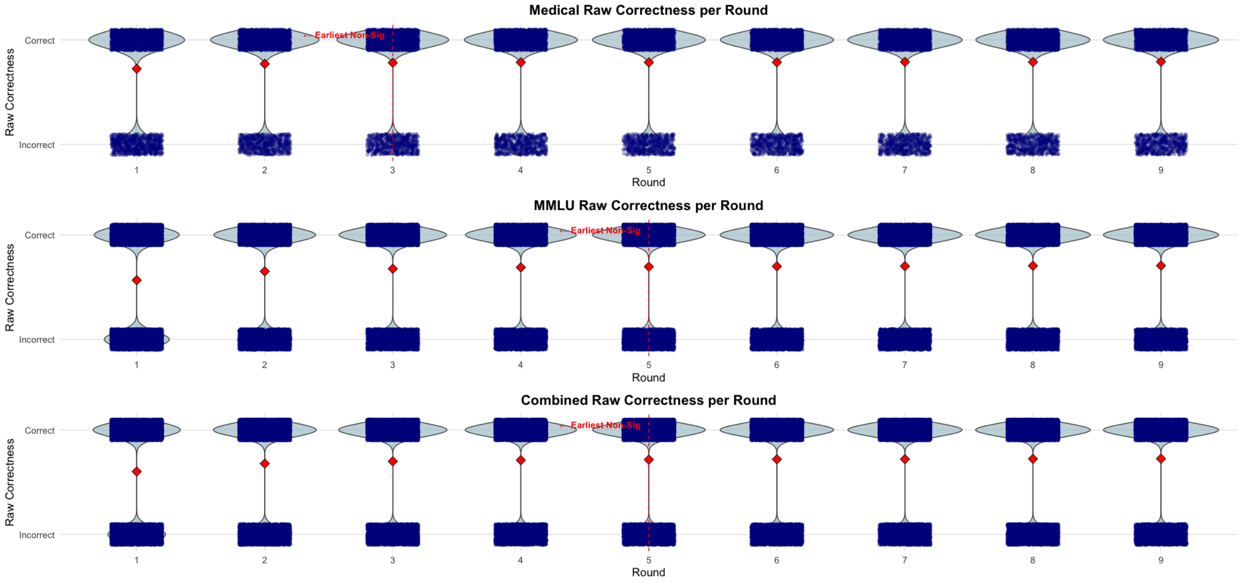
